# Diabetes mellitus mortality among elderly Brazilians, 2012-2022: an ecologic study

**DOI:** 10.1101/2025.11.26.25341115

**Authors:** Luciana Lima de Araújo, Thaiza Teixeira Xavier Nobre, Maria Eduarda Silva do Nascimento, Lidiany Galdino Felix, Richardson Augusto Rosendo, Ketyllem Tayanne da Silva Costa, Clemente Neves Sousa, Ana Elza Oliveira de Mendonça

## Abstract

**Objective:** This study aims to analyze the trend in the mortality rate due to Diabetes Mellitus among older adults in Brazil between 2012 and 2022.

**Methods:** An ecological study was conducted using secondary, publicly available data on deaths from DM (ICD-10 codes E10–E14) obtained from the Departamento de Informática do Sistema Único de Saúde (DATASUS). Standardized mortality rates were calculated based on the world standard population proposed by Segi (1960) and modified by Doll et al. (1966). Temporal trends were analyzed using the Prais-Winsten regression model, which corrects for first-order autocorrelation in time series. The Moran Global Index and Local Indicators of Spatial Association (LISA) were applied to assess spatial dependence and to identify high- and low-risk clusters across Brazilian regions.

**Results:** Between 2012 and 2022, Brazil recorded a total of [inserir número] deaths from DM among older adults. The standardized national mortality rate showed a decreasing trend during the period; however, regional disparities were evident. The North and Northeast regions presented higher and more persistent rates compared to the South and Southeast, which showed a more consistent decline. Spatial analysis revealed significant positive spatial autocorrelation (Moran’s I = [valor]; p < 0.05), identifying clusters of high mortality in municipalities of the Northeast and North regions.

**Conclusion:** The spatial and temporal analyses highlighted persistent regional inequalities in mortality due to DM in Brazil. While national mortality has declined, the concentration of high-risk clusters in socioeconomically vulnerable areas suggests that health interventions and policies must prioritize these regions. Understanding the spatial-temporal distribution of DM mortality contributes to more equitable public health planning, reinforcing the importance of targeted actions in prevention, early diagnosis, and access to continuous care for chronic conditions.

## Introduction

Noncommunicable chronic diseases (NCDs) are characterized as long-term conditions with persistent effects that can impact daily activities and require ongoing medical care. These diseases are considered the leading cause of death and disability worldwide, accounting for 71% of all causes of death globally. In Brazil, this proportion reaches 72%.^1^

Among these conditions is Diabetes Mellitus (DM) and its complications, which pose a significant and growing challenge to global public health. This disease is frequently associated with other clinical conditions such as hypertension, overweight, cardiovascular and kidney diseases, as well as with mortality due to COVID-19.^2^ In Brazil, it is estimated that DM is directly responsible for 6% of all deaths in the country and contributes to 31% of cardiovascular-related deaths.^3^

In this context, the research question guiding this study is: What is the trend in mortality due to Diabetes Mellitus in Brazil between 2012 and 2022? The relevance of this topic lies in the importance of discussing DM-related mortality in academic settings, as understanding this epidemiological profile may support the development of strategies and actions by policymakers and health professionals aimed at reducing these deaths, which are already considered preventable.

Therefore, this study aims to analyze the trend in the mortality rate due to Diabetes Mellitus among older adults in Brazil between 2012 and 2022.

## Methods

### Study design

An ecological study of spatial analysis and trend study of mortality from diabetes mellitus (DM) was conducted among the Brazilian population between the years 2012 and 2022. Data collection was carried out in April 2024, from DATASUS, in the Hospital Information System of the Unified Health System(SIH). Demographic information was extracted from the Brazilian Institute of Geography and Statistics (IBGE).

### Variables

As dependent variables, the study analyzed the mortality rate and the fatality rate due to DM. The independent variables consist of the number of hospitalizations and deaths due to DM, as well as the Brazilian states and years studied.

### Inclusion and exclusion criteria

All Brazilian federative units (26 states and the Federal District) with available and consistent data on mortality and cases of Diabetes Mellitus (DM) for the period from 2012 to 2022 were included in the study. The entire study was conducted with data only from elderly people (>60 years old). Exclusion criteria comprised unavailable, incomplete, or inconsistent data, as well as records with evidence of significant underreporting that could compromise the temporal analysis. Data from individuals under 60 years of age were excluded.

### Statistical analysis

Descriptive analysis was used to outline the profile of the sample used in this study. This analysis was presented through graphs and tables. The Moran Index (I) was calculated to identify evidence of spatial correlation in the data; furthermore, the Moran scatter plot was used to investigate the region in more detail.

For the calculation of mortality rates, population data from IBGE for the year 2022 were considered, and to calculate the fatality rate of the disease, the number of hospitalizations recorded in SIH/SUS during the study period was taken into account.

The standardized rates were calculated based on the standard world population proposed by Segi (1960) and later modified by Doll et al. (1966), according to the method adopted by the Cancer Incidence in Five Continents project of the International Agency for Research on Cancer (IARC).

The standardized specific mortality and case-fatality rates were obtained using the following mathematical expressions:

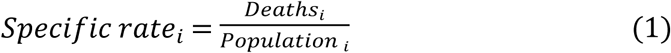

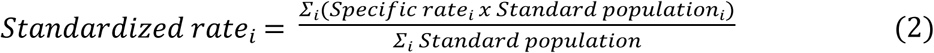

Where Population corresponds to the municipality’s population and Deaths corresponds to the number of hospitalizations or deaths from Diabetes Mellitus (DM) recorded in that municipality. The resulting standardized rate was multiplied by 100,000 for mortality rate estimates and by 100 for case fatality rate estimates.

For time trend analysis, the Prais-Winsten Regression method was used, suitable for models with first-order autocorrelation in the residuals (AR(1)). This method is widely recommended for time series studies in epidemiology, as it corrects for serial autocorrelation, ensuring greater precision and reliability in the estimates. This approach can be applied to different periods, as long as they are analyzed independently or comparatively, depending on the research objectives.

The robustness of the estimates was ensured by including an adequate number of temporal observations, respecting the minimum requirement of eight years recommended for this type of analysis. In the present study, the period covered 11 years (2012 to 2022), meeting the assumptions of regularity of time intervals and verification of residual autocorrelation, which reinforces the consistency of the results obtained.

For the trend analysis, Prais-Winsten regression was used. A logarithmic transformation was applied to the model in order to obtain the estimates of *β*1 for the number of deaths by state (3).

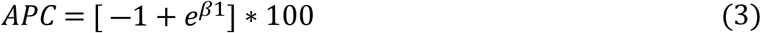

If APC > 1, then it can be said that the trend is increasing; if APC < 1, then it can be said that the trend is decreasing; and if APC = 0, or when there is no statistically significant difference from its value, that is, p-value > 0.05, then the trend is stationary. The analyses presented were conducted using the R programming language.

### Ethical review

The research using publicly available secondary data, provided in a completely anonymous manner, did not require approval from an Ethics Committee for research involving human subjects, in accordance with Resolution No. 510/2016 of the National Health Council.^4^

## Results

Figure 1 and Table 1 contain information regarding the mortality rate by state. The state with the highest mortality rate was in the state of Roraima, with a rate of 4.74. The lowest mortality rate was in the state of Paraná, with a rate of 0.85.

**Fig 1.**
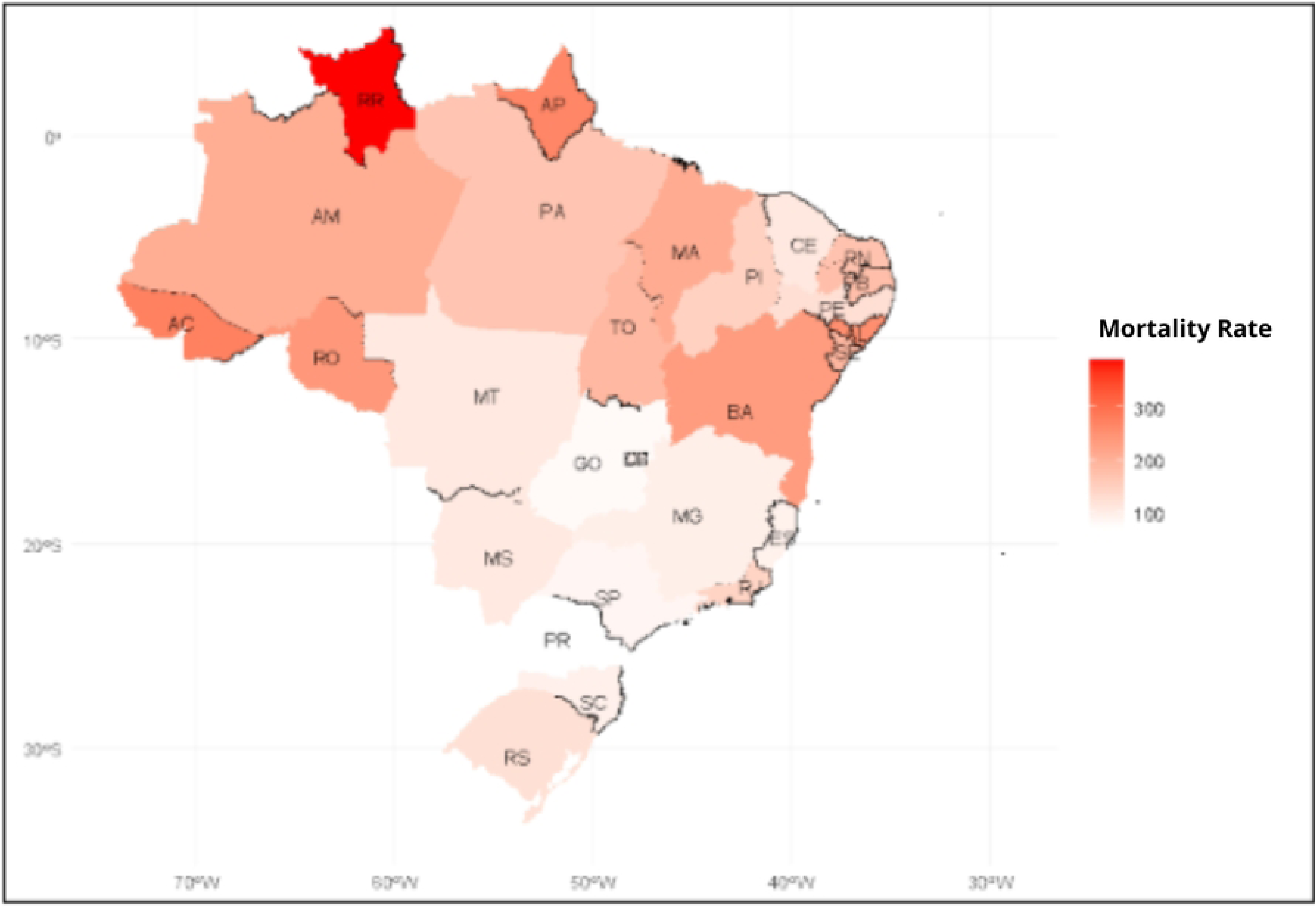
Diabetes mellitus mortality rate according to the Brazilian state. Brazil, 2024.

**Table 1.** Diabetes mellitus mortality rate according to the Brazilian state. Brazil, 2024.

| State | Total of deaths (%) | Mortality Rate |
| --- | --- | --- |
| Rondônia | 566 (1.2%) | 2.89 |
| Acre | 249 (0.5%) | 3.18 |
| Amazonas | 885 (1.8%) | 2.48 |
| Roraima | 239 (0.5%) | 4.74 |
| Pará | 1,776 (3.6%) | 2.03 |
| Amapá | 187 (0.4%) | 3.02 |
| Tocantins | 436 (0.9%) | 2.31 |
| Maranhão | 2,122 (4.3%) | 2.58 |
| Piauí | 906 (1.8%) | 1.83 |
| Ceará | 1,893 (3.9%) | 1.47 |
| Rio Grande do Norte | 1,112 (2.3%) | 2.23 |
| Paraíba | 1,430 (2.9%) | 2.32 |
| Pernambuco | 2,118 (4.3%) | 1.59 |
| Alagoas | 1,276 (2.6%) | 3.12 |
| Sergipe | 688 (1.4%) | 2.34 |
| Bahia | 6,419 (13.1%) | 2.97 |
| Minas Gerais | 4,436 (9.0%) | 1.21 |
| Espírito Santo | 725 (1.5%) | 1.15 |
| Rio de Janeiro | 5,206 (10.6%) | 1.72 |
| São Paulo | 7,829 (15.9%) | 1.02 |
| Paraná | 1,611 (3.3%) | 0.85 |
| Santa Catarina | 1,389 (2.8%) | 1.17 |
| Rio Grande do Sul | 3,251 (6.6%) | 1.48 |
| Mato Grosso do Sul | 505 (1.0%) | 1.29 |
| Mato Grosso | 545 (1.1%) | 1.27 |
| Goiás | 977 (2.0%) | 1.01 |
| Distrito Federal | 358 (0.7%) | 0.98 |
| **TOTAL** | **49,134** | **1.53** |

The Augmented Dickey-Fuller (ADF) test was performed to verify the hypothesis of stationarity of the time series (p-value = 0.01). Considering a significance level of 5%, the test indicates sufficient evidence to reject the null hypothesis, that is, the hypothesis that the series is not stationary. Therefore, it is concluded that the adjustment of a model for stationary series is appropriate for this data set. Next, the Moran index (I) was calculated to assess the presence of spatial autocorrelation in the data, obtaining I = 0.3873 (p-value = 0.0013). Given these results, there is statistically significant evidence of positive spatial correlation, indicating that spatially close observations tend to present similar values, as illustrated in Figure 1.

Figure 1 shows the spatial distribution of diabetes mellitus (DM) mortality rates across Brazil. Regional heterogeneity is evident, with higher mortality rates in the North and Northeast regions, especially in the states of Roraima (RR), Amazonas (AM), Acre (AC), and Bahia (BA), which are highlighted by darker tones on the map. These higher values suggest a greater burden of the disease in these areas, possibly associated with socioeconomic inequalities, reduced access to health services, and difficulties in continuous DM treatment monitoring.

In contrast, states in the Central-West, Southeast, and South regions—such as Goiás (GO), Minas Gerais (MG), Paraná (PR), and Santa Catarina (SC)—have lower adjusted rates, reflecting better primary care and health surveillance infrastructure, as well as greater coverage of chronic disease control programs. The presence of similar spatial patterns between neighboring states, evidenced by the continuous color gradient, reinforces the Moran index result (I = 0.3873; p = 0.0013), indicating positive spatial autocorrelation. This means that geographically close areas tend to have similar DM mortality rates, suggesting that shared regional factors, such as healthcare infrastructure, socioeconomic conditions, and dietary habits, directly influence mortality.

Overall, Figure 1 highlights the need for targeted public policies to reduce regional inequalities, with a special focus on the North and Northeast regions, where the burden of diabetes mortality remains most pronounced.

Figure 2 presents the Moran scatter plot, which presents a value of I = 0.3868 (p-value 0.0013). Considering the calculated values related to the Moran index, evidence of spatial correlation was found in the data. States in the first quadrant have a high mortality rate and are surrounded by states with high mortality rates. Sergipe and Roraima were located in the first quadrant. This indicates that Sergipe and Roraima exhibited a high mortality rate, which may be characterized by the region, considering that neighboring states also had high mortality rates.

**Fig 2.**
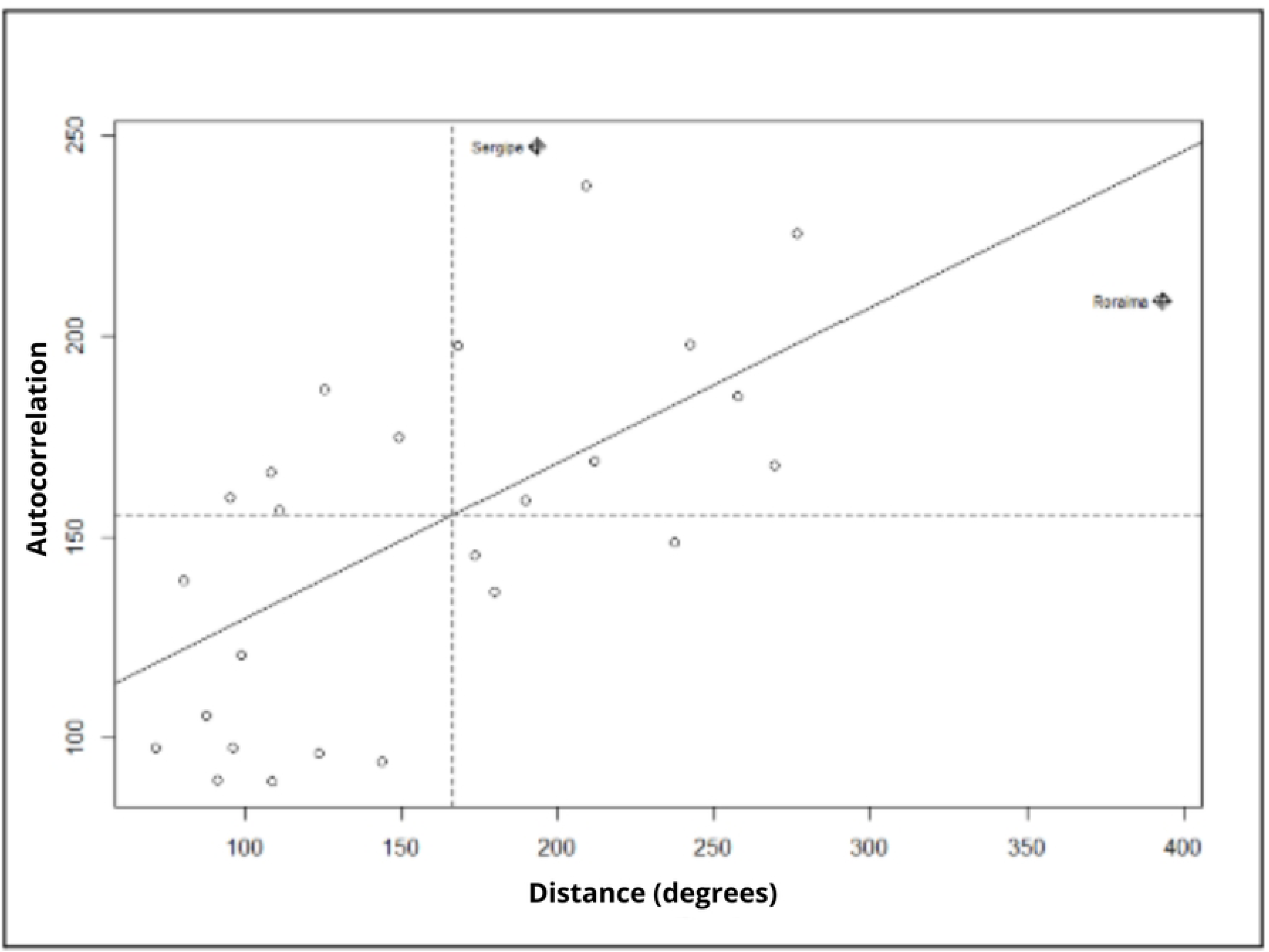
The Moran’s scatter plot. Brazil, 2024.

Figure 2 presents a Moran scatterplot, used to identify patterns of spatial autocorrelation between diabetes mellitus (DM) mortality rates in Brazilian states. The graph’s interpretation is based on the states’ position in relation to the quadrants: those located in the first quadrant (high-high) indicate high mortality rates associated with neighboring states that also have high values, constituting spatial clusters of high risk.

The states of Sergipe and Roraima were found to be located in this first quadrant, demonstrating a spatial concentration of high mortality rates. This behavior suggests the influence of shared regional factors, possibly related to the structure of health services, local socioeconomic conditions, and the effectiveness of diabetes prevention and control policies, which contribute to the maintenance of similar mortality patterns among contiguous states.

Table 2 and Figure 3 present the case fatality rate for diabetes mellitus in Brazil between 2012 and 2022. The Moran index presented a value of I = 0.1472 (p-value 0.1012). Considering the calculated values related to the Moran index, no evidence of spatial correlation was found in the data. The highest case fatality rate was found in the state of Rio de Janeiro, with 11.8 deaths for every 100 hospitalizations, while the lowest case fatality rate was in Paraná, with 3.53 deaths for every 100 hospitalizations.

**Fig 3.**
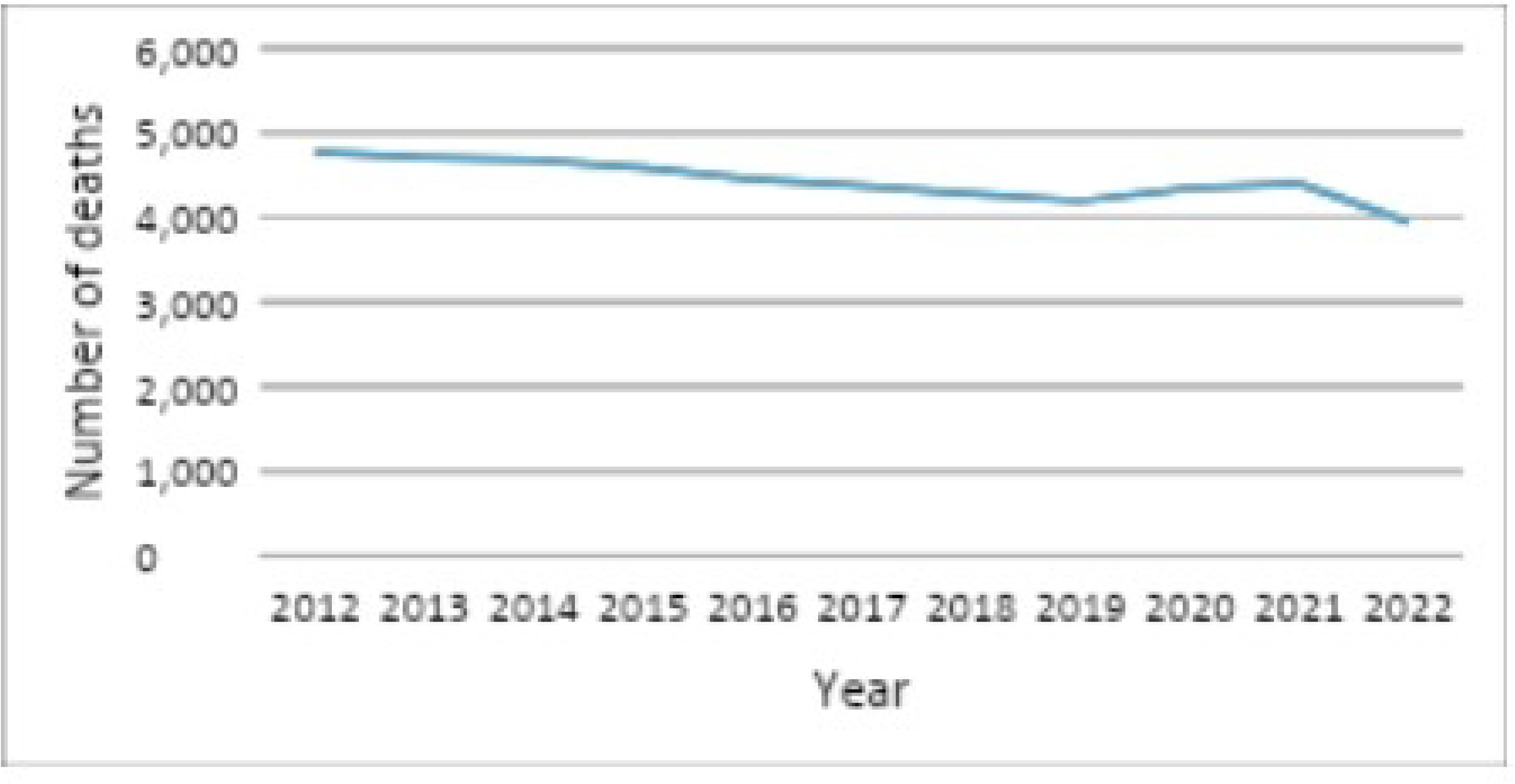
Diabetes mellitus fatality rate in Brazil. Brazil, 2024.

**Table 2.** Fatality rate by Brazilian state. Brazil, 2024.

| State | Hospitalizations | Deaths | Fatality rate |
| --- | --- | --- | --- |
| Rondônia | 11,992 (1.6%) | 566 (1.2%) | 4.72 |
| Acre | 2,586 (0.3%) | 249 (0.5%) | 9.63 |
| Amazonas | 15,212 (1.9%) | 885 (1.8%) | 5.82 |
| Roraima | 2,941 (0.4%) | 239 (0.5%) | 8.13 |
| Pará | 37,256 (4.7%) | 1,776 (3.6%) | 4.77 |
| Amapá | 1,861 (0.2%) | 187 (0.4%) | 10.05 |
| Tocantins | 6,905 (0.9%) | 436 (0.9%) | 6.31 |
| Maranhão | 56,960 (7.2%) | 2,122 (4.3%) | 3.73 |
| Piauí | 23,668 (3.0%) | 906 (1.8%) | 3.83 |
| Ceará | 29,811 (3.8%) | 1,893 (3.9%) | 6.35 |
| Rio Grande do Norte | 15,925 (2.0%) | 1,112 (2.3%) | 6.98 |
| Paraíba | 16,969 (2.1%) | 1,430 (2.9%) | 8.43 |
| Pernambuco | 35,836 (4.5%) | 2,118 (4.3%) | 5.91 |
| Alagoas | 12,435 (1.6%) | 1,276 (2.6%) | 10.26 |
| Sergipe | 6,193 (0.8%) | 688 (1.4%) | 11.11 |
| Bahia | 80,263 (10.2%) | 6,419 (13.1%) | 8.00 |
| Minas Gerais | 90,046 (11.4%) | 4,436 (9.0%) | 4.93 |
| Espírito Santo | 13,680 (1.7%) | 725 (1.5%) | 5.30 |
| Rio de Janeiro | 44,105 (5.6%) | 5,206 (10.6%) | 11.80 |
| São Paulo | 110,272 (14.0%) | 7,829 (15.9%) | 7.10 |
| Paraná | 45,580 (5.8%) | 1,611 (3.3%) | 3.53 |
| Santa Catarina | 25,387 (3.2%) | 1,389 (2.8%) | 5.47 |
| Rio Grande do Sul | 53,227 (6.7%) | 3,251 (6.6%) | 6.11 |
| Mato Grosso do Sul | 9,912 (1.3%) | 505 (1.0%) | 5.09 |
| Mato Grosso | 11,878 (1.5%) | 545 (1.1%) | 4.59 |
| Goiás | 21,620 (2.7%) | 977 (2.0%) | 4.52 |
| Distrito Federal | 6,941 (0.9%) | 358 (0.7%) | 5.16 |
| Total | 789,461 | 49134 | 6.22 |

The historical data show a downward trend in the number of deaths over the analyzed period. In 2012, 4,771 deaths were recorded, while in 2022 the number dropped to 3,944, representing an approximate decrease of 17.3%. This decrease may be related to increased access to pharmacological treatments, improvements in primary health care, and greater awareness of glycemic control and prevention of complications.

However, a stabilization and slight increase was observed between 2019 and 2021, a period that coincides with the COVID-19 pandemic, which significantly impacted the monitoring of chronic diseases and the functioning of health services. The resumption of the downward trend in 2022 suggests a gradual reorganization of the health system and the return of monitoring and prevention actions.

In the annual trend analysis, it is observed that, in Brazil, there are a total of 32,113,490 cases of diabetes mellitus and, from 2012 to 2022, there were a total of 49,134 deaths, with the highest fatality rate occurring in 2020.

**Table 3.** Number of deaths from DM in Brazil between 2012 and 2022 and fatality rate per year according to age group. Brazil, 2024.

| Year | 60-69 years | 70-79 years | 80 years and old | Total | Fatality Rate |
| --- | --- | --- | --- | --- | --- |
| 2012 | 1,459 | 1,731 | 1,581 | 4,771 | 6.2 |
| 2013 | 1,438 | 1,668 | 1,606 | 4,712 | 6.2 |
| 2014 | 1,383 | 1,663 | 1,633 | 4,679 | 6.2 |
| 2015 | 1,407 | 1,616 | 1,558 | 4,581 | 6.1 |
| 2016 | 1,354 | 1,601 | 1,501 | 4,456 | 6.4 |
| 2017 | 1,351 | 1,558 | 1,457 | 4,366 | 6.1 |
| 2018 | 1,356 | 1,524 | 1,403 | 4,283 | 6.0 |
| 2019 | 1,300 | 1,535 | 1,357 | 4,192 | 5.9 |
| 2020 | 1,409 | 1,560 | 1,373 | 4,342 | 6.8 |
| 2021 | 1,435 | 1,535 | 1,430 | 4,400 | 6.6 |
| 2022 | 1,176 | 1,442 | 1,326 | 3,944 | 6.0 |
| Total | 15,187 | 17,585 | 16,362 | 49,134 | 6.3 |

From 2012 to 2022, there was a general downward trend in the number of deaths from diabetes mellitus (DM) among the elderly in the age groups of 60 to 69 years, 70 to 79 years, and 80 years or older. Over the decade as a whole, 49,134 deaths were recorded, of which 15,187 were among people aged 60 to 69 years (30.9% of the total), 17,585 were among those aged 70 to 79 years (35.8%), and 16,362 were among those aged 80 years or older (33.3%).

These data indicate that, although all age groups showed a reduction over time, the 70 to 79 age group accounted for the highest proportion of deaths, remaining the most affected throughout the analyzed period. Overall, the total annual number of deaths fell from 4,771 in 2012 to 3,944 in 2022, representing a reduction of approximately 17.3% over ten years.

This decrease is similar across all age groups, but is most significant among individuals aged 60 to 69, who saw a drop from 1,459 deaths in 2012 to 1,176 in 2022, representing a reduction of 19.4%. The 70 to 79 age group dropped from 1,731 to 1,442 deaths (a 16.7% decrease), while among those over 80, the decrease was from 1,581 to 1,326 (16.1%). This downward trend corresponds to an average annual rate of decline (CAGR) of between -1.8% and -2.1%, demonstrating a gradual and consistent improvement in DM mortality indicators throughout the decade.

Despite the overall decline, there are specific fluctuations worth noting. The period from 2020 to 2021 saw a slight increase in total deaths, breaking the downward trend observed until 2019. The total fell from 4,192 in 2019 to 4,342 in 2020 and 4,400 in 2021, before falling again to 3,944 in 2022.

This increase coincides with the context of the COVID-19 pandemic, which directly impacted the management of chronic diseases. The overload on health services, the interruption of regular follow-up care, and the greater vulnerability of people with diabetes to infection may have contributed to the increase in deaths in these two years.

In 2022, a sharp decline was noted, especially among individuals aged 60 to 69, who recorded an 18.1% reduction compared to 2021. This may indicate a partial recovery in health services and clinical monitoring, although it may also be related to possible variations in reporting or delays in notifications.

The analysis of the case fatality rate, measured per 100 hospitalizations, reinforces the relative stability of the indicators over time. During the period analyzed, this rate ranged from 5.9 (in 2019) to 6.8 (in 2020), with an overall average of 6.3. The peak in 2020, again, coincides with the most critical period of the pandemic, when there was greater severity of hospitalized cases and difficulties in accessing hospital care.

After this increase, the case fatality rate gradually declined, returning to 6.0 in 2022, a value similar to that observed before the health crisis. The observed annual fluctuations are moderate, indicating a solid downward trend, albeit subject to variations due to external factors such as health crises and changes in health policies.

The continued decline suggests significant advances in the diagnosis, treatment, and control of diabetes mellitus, possibly associated with public primary care policies and expanded access to medication and regular monitoring. The continued high concentration of deaths in the 70- to 79-year-old age group reinforces the need for specific prevention and care strategies for this group, which combines a high prevalence of the disease with physiological vulnerabilities associated with aging.

On the other hand, the abrupt decrease in deaths in 2022 requires cautious interpretation. It is important to determine whether this decline reflects an effective improvement in health conditions and diabetes control or is due to administrative factors such as delays in consolidating records or changes in the coding of causes of death.

Epidemiologically, the scenario reveals a positive evolution, marked by a sustained reduction in DM deaths among the elderly, with the exception of the temporary impact observed during the pandemic. The stability of the fatality rate suggests that, even with fluctuations in the absolute number of deaths, the average severity of hospitalized cases remained constant. However, the lack of population data by age group limits the analysis of specific mortality rates, preventing standardization per 100,000 population and direct comparison between years and regions.

Table 4 presents the annual variation (%), p-value, and mortality trend by Brazilian state. Brazil exhibited a decreasing trend in mortality due to DM over the years, with a variation of -1.48%.

**Table 4.** Analysis of mortality trends by the Brazilian state. Brazil, 2024.

| State | Annual variation (%) | p-value | Tendency |
| --- | --- | --- | --- |
| Brasil | -1.48 | **<0.001** | Decreasing |
| Rondônia | -1.01 | 0.38 | Stationary |
| Acre | -0.30 | 0.751 | Stationary |
| Amazonas | 11.88 | **0.001** | Increasing |
| Roraima | 3.12 | 0.459 | Stationary |
| Pará | 1.04 | 0.363 | Stationary |
| Amapá | -2.67 | 0.264 | Stationary |
| Tocantins | -4.95 | **<0.001** | Decreasing |
| Maranhão | 3.25 | **0.048** | Increasing |
| Piauí | -2.15 | 0.182 | Stationary |
| Ceará | -1.69 | 0.148 | Stationary |
| Rio Grande do Norte | -4.60 | **<0.001** | Decreasing |
| Paraíba | -2.86 | **0.008** | Decreasing |
| Pernambuco | -7.46 | **<0.001** | Decreasing |
| Alagoas | -10.72 | **<0.001** | Decreasing |
| Sergipe | 0.59 | 0.863 | Stationary |
| Bahia | -1.13 | 0.113 | Stationary |
| Minas Gerais | 0.88 | 0.338 | Stationary |
| Espírito Santo | -0.10 | 0.941 | Stationary |
| Rio de Janeiro | -3.84 | **<0.001** | Decreasing |
| São Paulo | -1.11 | **0.012** | Decreasing |
| Paraná | -0.45 | 0.683 | Stationary |
| Santa Catarina | 1.10 | 0.054 | Stationary |
| Rio Grande do Sul | -1.60 | 0.054 | Stationary |
| Mato Grosso do Sul | -0.20 | 0.927 | Stationary |
| Mato Grosso | 0.62 | 0.63 | Stationary |
| Goiás | -1.70 | 0.201 | Stationary |
| Distrito Federal | -12.79 | **<0.001** | Decreasing |

Analysis of the data presented in Table 4 reveals distinct patterns in the annual variation in mortality rates from Diabetes Mellitus (DM) among Brazilian states during the period evaluated. At the national level, a significant downward trend is observed (−1.48%; p < 0.001), indicating that, nationwide, there has been a consistent reduction in mortality rates associated with the disease over the years.

When analyzing the individual states, it was found that some states showed statistically significant downward trends, notably Tocantins (−4.95%; p < 0.001), Rio Grande do Norte (−4.60%; p < 0.001), Pernambuco (−7.46%; p < 0.001), Alagoas (−10.72%; p < 0.001), Rio de Janeiro (−3.84%; p < 0.001), São Paulo (−1.11%; p = 0.012), and the Federal District (−12.79%; p < 0.001). These results suggest significant advances in diabetes prevention and management, possibly associated with strengthening primary care, expanding public policies, and improving access to medications and ongoing care.

On the other hand, some states showed an upward trend in mortality rates, notably Amazonas, with a significant annual growth of 11.88% (p = 0.001), and Maranhão, with 3.25% (p = 0.048). These findings indicate a worsening of the local epidemiological situation, which may reflect limitations in health service coverage, regional inequalities, or difficulties in accessing diagnosis and appropriate treatment.

Most of the remaining states showed a steady trend, with no statistically significant variations, indicating stability in mortality rates, albeit at different levels. This stability may be associated with both the maintenance of control policies and the persistence of structural factors that impede more significant progress.

Overall, the results point to regional heterogeneity in the behavior of DM mortality in Brazil. While regions such as the Northeast and Southeast show significant progress, the North Region, especially Amazonas, highlights persistent challenges in combating the disease, reinforcing the need for targeted interventions and more equitable health policies among Brazilian states.

## Discussion

The results demonstrated a general downward trend in diabetes mellitus (DM) mortality in Brazil between 2012 and 2022, with an approximate 17.3% decline in the total number of deaths recorded during this period. This reduction reflects the expansion of prevention and control efforts for chronic noncommunicable diseases (NCDs) in the country, reflecting advances in public health policies, expanded access to medication, and the consolidation of primary care as the gateway to the Unified Health System (SUS).^1,10^

The gradual decline between 2012 and 2018 can be attributed to the effectiveness of programs focused on glycemic control, early diagnosis, and health education, which is consistent with evidence highlighting the role of healthy lifestyle pillars in managing NCDs.^1^

However, temporal analysis revealed a specific increase in mortality rates between 2020 and 2021, especially in older age groups. This increase coincides with the COVID-19 pandemic, which caused disruptions in outpatient care, difficulties in accessing health services, and worsening clinical conditions for individuals with pre-existing chronic conditions.^13,15^

Research has shown that diabetes was one of the main factors associated with in-hospital mortality from COVID-19 in Brazil, both due to increased immunological vulnerability and reduced clinical monitoring capacity during the pandemic.^13,14^ Nevertheless, the downward trend resumed in 2022, suggesting a reorganization of the health system and the resumption of monitoring and prevention actions.^15^

Spatial analysis revealed significant positive autocorrelation (Moran’s I index = 0.3873; p = 0.0013), indicating that neighboring states tend to exhibit similar DM mortality patterns. This heterogeneous spatial distribution revealed a higher concentration of deaths in the North and Northeast regions, especially in the states of Roraima, Amazonas, Acre, and Bahia, while the lowest adjusted rates were observed in the South, Southeast, and Central-West regions, such as Goiás, Minas Gerais, Paraná, and Santa Catarina. These results corroborate previous findings that indicate a direct association between human development indicators and mortality from NCDs in Brazil.^2,11^

Studies show that regions with lower Human Development Index (HDI) and more limited primary care coverage are more vulnerable to DM mortality, resulting from socioeconomic inequalities, difficulties in accessing supplies and medications, and a lower capacity for longitudinal patient monitoring.^2,11,17^

Conversely, regional economic development can exert a dual influence on the epidemiology of diabetes. On the one hand, socioeconomic growth and the expansion of public health networks tend to reduce mortality rates by improving early diagnosis, treatment adherence, and clinical management.^9,10^

On the other hand, urbanization and cultural changes resulting from development may contribute to an increase in the incidence of the disease, due to the adoption of high-calorie diets, sedentary lifestyles, and population aging.^11,16^ Therefore, the observed reduction in mortality should not be interpreted in isolation, but rather in the context of social and cultural transformations that affect the standard of living and health determinants of the Brazilian population

Regarding age distribution, the expected pattern of higher mortality among older age groups remained, indicating that the risk of death from DM increases progressively with aging. This result is consistent with the literature, which identifies aging as one of the main vulnerability factors for complications and fatal outcomes associated with diabetes.^8,12^

Inadequate glycemic control, the presence of comorbidities, and the accumulation of metabolic damage over the years contribute to worsening prognosis in the elderly.^13,16^ The maintenance of this age gradient reinforces the need for continuous care policies aimed at this population, especially in regions where access to specialized services is limited.

The identified spatial heterogeneity reinforces the importance of coping strategies adapted to regional realities. In the North and Northeast, strengthening Primary Health Care (PHC) is essential, with an emphasis on screening, nutritional education, and the control of risk factors such as obesity and sedentary lifestyles. In more developed regions, where access to services is broader, the priority should be improving health surveillance, integrating information systems, and reducing intraregional inequalities. Successful experiences, such as the use of risk stratification technologies in diabetic patients^6^ and comprehensive diabetic foot care programs, show promise for reducing complications and hospitalizations, indirectly impacting mortality rates.

Despite the advances, some limitations should be considered. The use of secondary mortality data is subject to underreporting and recording delays, estimated at up to two years in some databases, which may underestimate the most recent figures and influence the observed temporal trends.^4^ Furthermore, there are regional variations in the accuracy of the underlying cause of death, resulting from differences in diagnostic capacity and the quality of death certificate completion.^12^

The ecological design employed limits causal inferences at the individual level, since the observed associations between regional variables and mortality do not necessarily reflect individual behaviors.^5,7^ Finally, factors such as obesity prevalence, glycemic control, therapeutic adherence, and Family Health Program coverage were not directly included in the analysis and may act as confounding variables.^16^

Even given these limitations, the results as a whole demonstrate significant progress in reducing DM mortality in Brazil over the last decade, partially reflecting the positive impact of public policies focused on NCDs. However, the observed regional disparities indicate that progress has not been homogeneous and that structural inequalities in access to health care persist. Effectively combating diabetes requires integrated actions across levels of care, strengthening epidemiological surveillance, and expanding intersectoral policies that address the social determinants of health. Therefore, the continuity and improvement of national strategies, especially in the North and Northeast regions, are fundamental to consolidate the advances achieved and reduce inequities in mortality from Diabetes Mellitus in Brazil.

## Conclusion

The temporal and spatial analysis of mortality due to Diabetes Mellitus (DM) in Brazil between 2012 and 2022 revealed a heterogeneous pattern across regions. Although there was a general decline in mortality rates nationwide, the North and Northeast regions still showed higher and more persistent rates, indicating slower progress compared to the South and Southeast. These disparities suggest that regional inequalities in access to healthcare, early diagnosis, and continuous treatment remain key determinants of mortality from DM.

The temporal trend indicates improvements in disease management and greater effectiveness of public health policies in certain regions, while the spatial concentration of higher mortality rates in areas with lower socioeconomic indicators highlights the need for targeted interventions. Strengthening primary healthcare, expanding preventive programs, and improving the monitoring of chronic diseases are essential strategies to reduce these regional disparities.

Therefore, understanding the spatial-temporal distribution of DM mortality allows for more precise decision-making by health managers and supports the formulation of public policies focused on equity, aiming to ensure access to prevention and treatment resources in all Brazilian regions.

## Data Availability

All data files are available from the DATASUS database (Link: https://datasus.saude.gov.br/informacoes-de-saude-tabnet/).

## Financing

This study will be partially funded by the Coordination for the Improvement of Higher Education Personnel-Brazil (CAPES): Financial Code 001. Funders will have no role in the study design, data collection and analysis, publication decision or preparation of the manuscript.

